# Commercial stocks of SARS-CoV-2 RNA may report low concentration values, leading to artificially increased apparent sensitivity of diagnostic assays

**DOI:** 10.1101/2020.04.28.20077602

**Authors:** Erik Jue, Rustem F. Ismagilov

**Affiliations:** Division of Biology and Biological Engineering; Division of Chemistry and Chemical Engineering

## Abstract

In response to the rapidly evolving COVID-19 pandemic, the U.S. Food and Drug Administration (FDA) has rapidly issued 49 emergency use authorizations (EUAs) for SARS-CoV-2 in vitro diagnostic test-kits. A critical metric in the performance evaluation for a diagnostic test kit is the analytical sensitivity, which is measured by the limit of detection (LOD). Commercial RNA stocks with known titers are used to determine LOD. We identified a problem with the titer reported for the commercial stocks when examining the analytical sensitivity of the reverse transcription quantitative PCR (RT-qPCR) protocol that is recommended by the Centers for Disease Control and Prevention (CDC) using plasmid DNA from Integrated DNA Technologies (IDT), synthetic RNA from BEI Resources (BEI), and extracted genomic RNA from BEI. We detected 3/3 positives for reactions containing synthetic RNA at a concentration of 0.1 copies/reaction (based on the supplier’s label concentration). The apparent better-than-single-molecule performance is a statistically highly unlikely event, indicating a potential inaccuracy in the supplier’s quantification of the stock material. Using an ultrasensitive and precise assay, reverse transcription digital PCR (RT-dPCR), we independently quantified concentrations of commercial SARS-CoV-2 plasmid DNA and SARS-CoV-2 RNA stocks. For plasmid DNA, the actual concentration measured by RT-dPCR was 11% of the nominal label concentration. For synthetic RNA, the actual concentration measured by RT-dPCR for one lot was 770% of the label concentration and for a different lot was 57% of the label concentration. For genomic RNA, the concentration measured by RT-dPCR for one lot was 240% of the label concentration and for a different lot it was 300% of the label concentration. This SARS-CoV-2 genomic RNA from BEI Resources has been used in at least 11 approved FDA Emergency Use Authorizations as of April 27, 2020. Such deviations of reported RNA or DNA stock concentrations from true concentrations can result in inaccurate quantification and calculation of LOD. Precise and accurate reporting of DNA and RNA stock concentrations by commercial suppliers will enable accurate quantification of assay performance, which is urgently needed to improve evaluation of different assays by diagnostic developers and regulatory bodies.

## Introduction

As of April 27, 2020, the COVID-19 pandemic has reached 185 countries/regions, with more than 3 million infected individuals, and more than 210,000 deaths.[1, 2] The rapid spread of SARS-CoV-2 (the virus that causes COVID-19) and the large proportion of asymptomatic infected individuals has led to widespread demand for diagnostic test kits. To meet the massive demand, on February 4, 2020, the U.S. Department of Health and Human Services (HSS) secretary declared that circumstances exist justifying the authorization of emergency use of in vitro diagnostics for detecting SARS-CoV-2.[3] Since then, the U.S. Food and Drug Administration (FDA) has fast-tracked 49 Emergency Use Authorizations (EUAs) for SARS-CoV-2 test-kit manufacturers and commercial laboratories.[4]

The application to receive an EUA requires a description of the assay and an evaluation of its performance.[5] A key metric in evaluating assay performance is the analytical sensitivity, which describes the ability of a test to detect very low concentrations of the target analyte. Analytical sensitivity is typically measured using the limit-of-detection (LOD), which is the concentration of target analyte that can be consistently detected at least 95% of the time (19 of 20 replicates are positive). It is important for test kits to demonstrate a low LOD, which indicates good sensitivity of the test and the ability to detect samples containing very low viral RNA concentrations. Thus far, many diagnostic test kits for SARS-CoV-2 RNA have reported good LODs, with some reporting down to as low as 40 copies/mL.

We obtained SARS-nCoV-2 plasmid DNA from IDT, synthetic SARS-nCoV-2 RNA from BEI, and genomic RNA from SARS-nCoV-2, isolate USA-WA1/2020 from BEI. In a well-functioning assay (i.e., perfect transcription of RNA), we would expect to observe the same LOD for all 3 stocks. Instead, using the quantitative reverse transcription polymerase chain reaction (RT-qPCR) protocol recommended by the Centers for Disease Control and Prevention (CDC), we found substantial discrepancies, leading us to question the accuracy of the reported stock concentrations from the commercial suppliers.

In this manuscript, we aimed to resolve the discrepancies in the LODs obtained with the three SARS-CoV-2 NA stocks by performing our own quantification of each stock concentration using a highly sensitive digital quantification method, reverse transcription digital PCR (RT-dPCR).[6, 7]

## Results and Discussion

### Half-log dilutions of NA stocks in RT-qPCR

We first performed half-log dilutions on each SARS-CoV-2 NA stock (plasmid DNA from 10,000 to 3.16 nominal copies/reaction and synthetic RNA and genomic RNA from 100 to 0.0316 nominal copies/reaction) using the CDC-recommended RT-qPCR protocol (Fig. 1). A priori, we expect all NA stocks to have similar LOD. We also expect that the RNA LOD may be slightly worse than the DNA LOD because RNA must be transcribed prior to amplification. Contrary to these expectations, we observed that both RNA stocks outperformed the DNA stock. The first non-detects (reactions failing to amplify) appeared at 31.6 copies/reaction for plasmid DNA (Fig. 1A), at 0.0316 copies/reaction for synthetic RNA (Fig. 1B), and at 0.316 copies/reaction for genomic RNA (Fig. 1C). We also observed lower Cq values for synthetic RNA compared with genomic RNA at the same input dilution (using the supplier’s label concentration).

**Fig. 1:**
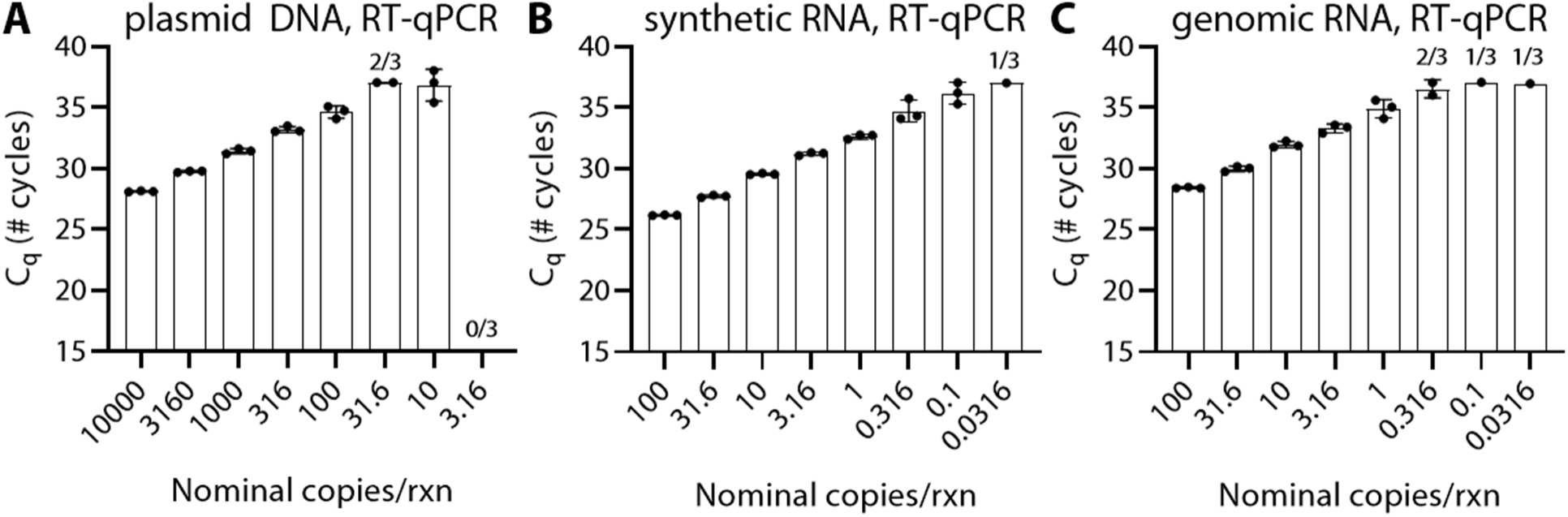
Better-than-statistically-likely performance of reverse-transcription quantitative PCR (RT-qPCR) protocol recommended by the CDC using half-log dilution series of SARS-CoV-2 nucleic-acid targets. Plots show number of quantitation cycles (C_q_) for half-log dilutions of (A) plasmid DNA lot 508728 from Integrated DNA Technologies (IDT), (B) synthetic RNA lot 70033953 from BEI Resources (BEI), and (C) genomic RNA lot 70033700 from BEI. No-template controls (n=12) did not amplify after 80 cycles.

Of particular interest, when quantifying the synthetic RNA, we detected 3/3 positives at a dilution of just 0.1 copies/reaction, indicating a virtually impossible better-than-single-molecule assay performance. An RT-qPCR reaction requires a minimum of 1 RNA copy as a template to exponentially amplify and generate a detectable signal. Better-than-single-molecule assay performance is highly unlikely based on statistics. Using the Poisson distribution,[8] which accounts for the stochasticity in loading a reaction well, when loading a solution into a well at an average target RNA concentration of 0.1 copies/reaction there is a ~9.5% chance that a reaction well actually contains at least one RNA copy. If testing a set of 3 reaction wells, the probability that all 3 wells contains at least one RNA copy each drops to 0.086% (0.095^3^).

### Re-quantification of NA stocks with RT-dPCR

To resolve the differences in LOD and understand the apparent better-than-single-molecule performance of the CDC assay, we re-quantified the NA stocks using an RT-dPCR protocol (Bio-Rad dPCR EvaGreen Supermix with added WarmStart Rtx (NEB) (Fig. 2). We first ran 16 no-template controls and measured an average background concentration of 1.9 ± 1.0 copies/µL. We defined the assay detection limit (99.7% confidence) as the background concentration plus 3 standard deviations of the background,[9] and calculated the assay detection limit of RT-dPCR to be 4.9 copies/µL. To quantify each stock concentration, we first diluted the NA stock down to a concentration within the digital quantification range (10 - 120,000 input copies of target) and measured with RT-dPCR.[10] We then took the concentration obtained from the RT-dPCR measurement, subtracted the background concentration, and multiplied the result by the dilution factor to calculate our RT-dPCR measured stock concentration.

**Fig. 2:**
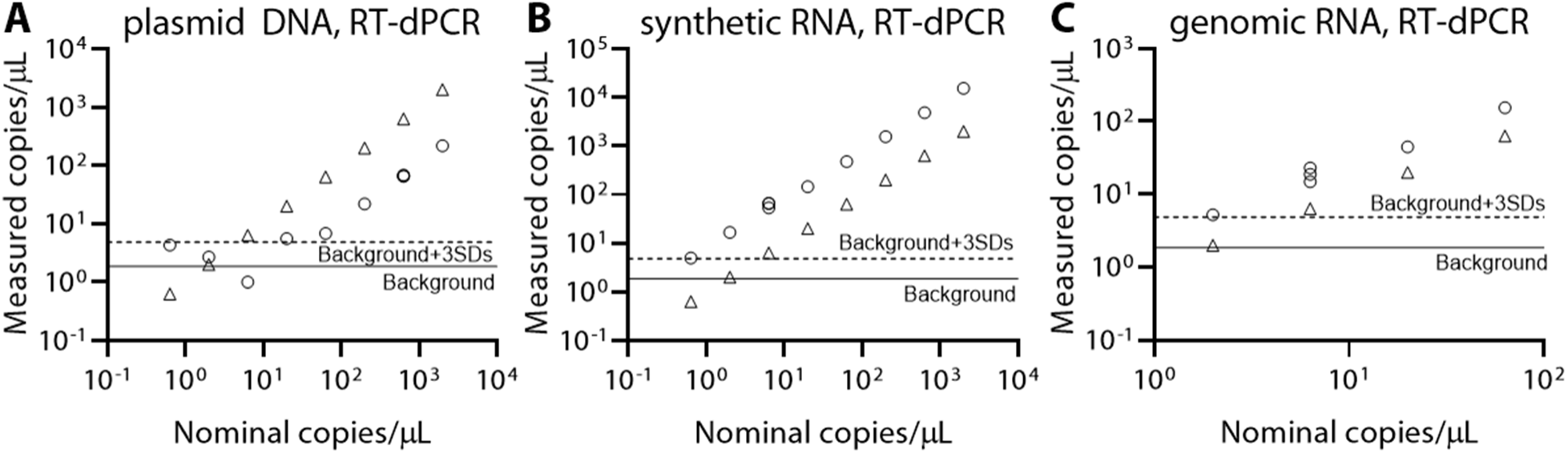
Quantification of different SARS-CoV-2 nucleic acid targets using an RT-dPCR protocol. Plots show measured concentration (circles) for half-log dilutions of the label concentrations (triangles) based on supplier-reported values of (A) plasmid DNA (lot 508728 from IDT), (B) synthetic RNA (lot 70033953 from BEI), and (C) genomic RNA (lot 70033700 from BEI). Individual dilution series were repeated twice more for the 633 copies/µL dilution with plasmid DNA and the 6.33 copies/µL dilution for both RNA stocks. The background (solid line) was calculated by averaging no-template controls (n=16), and the assay detection limit (dashed line) was calculated as the background signal in no-template controls plus 3 standard deviations of the background signal (99.7% confidence).[9]

We ran a dilution series for each SARS-CoV-2 NA stock. For plasmid DNA (IDT, lot 508528), we observed that all RT-dPCR quantifications were systematically lower than the concentration we expected from the supplier’s label concentration, except when our measurements dropped below the assay detection limit (Fig. 2A). To further validate our results, we performed 2 additional dilutions down to what should have been 633 copies/µL. Of the three measurements taken at 633 copies/µL, two were performed using RT-dPCR and the third was performed using dPCR (no reverse transcription). The RT-dPCR measurements were 69 and 66 copies/ µL and the dPCR measurement was 65 copies/µL. As expected for DNA, there was no difference in our measurements when using the dPCR assay with or without reverse transcription. To reduce the contributions of Poisson noise and background signal on our quantification, we used the highest concentration that we tested to quantify the stock concentration. Using the dilution for which we expected 2000 plasmid DNA copies/µL, we actually measured 220 copies/µL. From this measurement, we calculated a stock concentration of 2.2×10^4^ copies/µL, which is 11% of the supplier’s label concentration of 2×10^5^ copies/µL.

For synthetic RNA (BEI, lot 70033953), we observed that all RT-dPCR quantifications were systematically higher than the supplier’s label concentration of 5×10^5^ copies/µL (Fig. 2B). We performed two additional dilutions (three in total) down to what should have been 6.33 copies/µL and observed that all three values were higher than expected (54, 67, and 66 copies/µL). In a separate experiment, we performed dPCR (no reverse transcriptase) and observed that the signal was below background levels, which is expected because no amplification should occur in an RNA sample in the absence of reverse transcriptase. Using the dilution for which we expected 2000 copies/µL (based on the label concentration), we actually measured 15,000 copies/µL. From this measurement, we calculated a stock concentration of 3.9×10^6^ copies/µL, which is 770% of the supplier’s label concentration of 5×10^5^.

For genomic RNA (BEI, lot 70033700), we observed that all RT-dPCR quantifications were systematically higher than the supplier’s label concentration of 5.5×10^4^ copies/µL (Fig. 2C). We performed two additional dilutions (three in total) down to what should have been 6.33 copies/µL and observed that all three values were higher than expected (19, 15, and 23 copies/µL). The genomic RNA in a dPCR (no reverse transcription) experiment also measured below the background signal. Using the dilution for which we expected 63.3 copies/µL, we actually measured 155 copies/µL. From this measurement, we calculated a stock concentration of 1.3×10^5^ copies/µL, which is 240% of the supplier’s label concentration of 5.5×10^4^.

We next investigated potential differences among lots of stock SARS-CoV-2 RNA. We obtained an additional lot of synthetic RNA (lot 70034198) and an additional lot of genomic RNA (lot 70033320) from the same supplier (BEI). For each, we diluted the label concentration down to what should have been 63.3 copies/µL and measured the concentration with RT-dPCR (n=1). The synthetic RNA stock (lot 70034198) measured 38 copies/µL. From this, we calculated a concentration of 1.7×10^5^ copies/µL, which is 57% of the label concentration of 2.9×10^5^ copies/µL. We note that lot 70034198 was much closer (57%) to the label value as compared with lot 70033953 (770%). The genomic RNA stock (lot 70033320) measured 193 copies/µL. From this measurement, we calculated a concentration of 1.4×10^5^ copies/µL, which is 300% of the label concentration of 4.8×10^4^ copies/µL. We found that the RT-dPCR measurements for genomic RNA stocks were similarly higher (240% for lot 70033700, 300% for lot 70033320) than the supplier’s label concentration.

### Dilutions of NA stocks in RT-qPCR with RT-dPCR correction

Next, we selected concentrations near the LOD for each NA stock, and reran RT-qPCR with greater sample size (N=10 for each of three dilutions of each stock, Table 1). If we use the suppliers’ label concentrations to understand the RT-qPCR results, we observed large deviations (orders of magnitude differences) from expected values. For example, 3 of 10 wells turned positive for a label concentration diluted to 0.0316 copies/reaction with synthetic RNA (lot 70033953), whereas we would expect (based on statistical calculations) for 3 out of 100 wells to be positive. We also note that only 7 of 10 wells turned positive for a label concentration diluted to 31.6 copies/reaction with plasmid DNA (lot 528728), whereas statistically we expect all 10 wells to turn positive (assuming single-molecule detection). By Poisson distribution, we would expect 1 out of every 5×10^13^ wells to be negative at this concentration.

**Table 1:** Analytical performance near the limit of detection (LOD) for different SARS-CoV-2 targets in the CDC-recommended RT-qPCR protocol. Each nucleic acid stock was diluted and we selected 3 different concentrations to spike into RT-qPCR reaction wells (10 replicates for each condition). Measured concentrations were used to convert the label concentrations based on the RT-dPCR measurements reported in Fig. 2. Positives were counted as detected if they amplified within 40 cycles. No-template controls (N=6) did not amplify after 80 cycles.

|  | **Label**<br>**concentration**<br>**(copies/reaction)** | **RT-dPCR measured**<br>**concentration**<br>**(copies/reaction)** |
| --- | --- | --- |
| **plasmid DNA (IDT)**<br>**lot 508728** |  |  |
| Detected 7 of 10 | 31.6 | 3.45 |
| Detected 3 of 10 | 10 | 1.09 |
| Detected 2 of 10 | 3.16 | 0.345 |
| **synthetic RNA (BEI)**<br>**lot 70033953** |  |  |
| Detected 10 of 10 | 0.316 | 2.44 |
| Detected 8 of 10 | 0.1 | 0.772 |
| Detected 3 of 10 | 0.0316 | 0.244 |
| **genomic RNA (BEI)**<br>**lot 70033700** |  |  |
| Detected 9 of 10 | 1 | 2.42 |
| Detected 6 of 10 | 0.316 | 0.765 |
| Detected 5 of 10 | 0.1 | 0.242 |

If we instead use our RT-dPCR measured concentrations to understand our RT-qPCR results, the observed results are statistically more likely for all 3 NA stocks. The highest tested concentrations of plasmid DNA (lot 5208728), synthetic RNA (lot 70033953), and genomic RNA (lot 70033700) were measured by RT-dPCR to have concentrations of 3.45, 2.44, and 2.42 copies/reaction respectively. We observed 70%, 100%, and 90% positives for these conditions, which is a reasonable result considering that the Poisson distribution corresponding to 3 copies/reaction predicts an expected value (most likely) of 95% positives. For the tested concentrations measured by RT-dPCR to be near 1 copy/reaction with an expected value of 63% positives, we observed 30%, 80%, and 60% positives. Lastly, for the lowest tested concentrations measured by RT-dPCR to be near 0.3 copies/reaction with an expected value of 26% positives, we observed 20%, 30%, and 60% positives.

## Conclusions

In this manuscript, we observed vast discrepancies in the analytical sensitivity of the CDC-recommended RT-qPCR protocol based on which commercial NA stock was used. Performing an ultrasensitive digital PCR method, RT-dPCR, revealed there are likely errors in the supplier-reported concentrations. Specifically, using RT-dPCR, the measured concentration of plasmid DNA (IDT, lot 508728) was 11% of the label concentration. For synthetic RNA (BEI), the measured concentration by RT-dPCR was 770% of the label concentration for lot 70033953, whereas it was 57% of the label concentration for lot 70034198. For genomic RNA (BEI), the measured concentration by RT-dPCR was 240% of the label concentration for lot 70033700 and 300% of the label concentration for lot 70033320. An underreporting of the stock concentration could lead to an artificially improved LOD for a diagnostic assay. Such discrepancies in how NA suppliers quantify their stock can introduce significant biases and impair proper development and evaluation of in vitro diagnostics being considered for regulatory approvals and mass production.

The inaccurate quantification of the SARS-nCoV-2 genomic RNA concentration is concerning because our analysis of EUA documents indicated that this RNA stock (BEI, NR-52285) has been used in at least 11 EUAs: Lyra SARS-CoV-2 Assay, Abbott RealTime SARS-CoV-2 assay, AvellinoCoV2 test, NxTAG CoV Extended Panel Assay, NeuMoDx SARS-CoV-2 Assay, COV-19 IDx assay, BioGX SARS-CoV-2 Reagents for BD MAX System, ARIES SARS-CoV-2 Assay, Logix Smart Coronavirus Disease 2019 (COVID-19) Kit, Smart Detect SARS-CoV-2 rRT-PCR Kit, BD SARS-CoV-2Reagents for BD MAX System, and Curative-Korva SARS-Cov-2 Assay.[4] We note that the GeneFinder COVID-19 Plus RealAmp Kit and the STANDARD M nCoV Real-Time Detection Kit both report using genomic RNA, but we could not determine whether or not BEI was the supplier. Of the kits that used BEI genomic RNA, the majority did not report the lot or starting stock concentration (which could be used to deduce the lot number).

Discrepancies in stock quantification can be attributed to multiple factors, including differences in quantification method, reaction conditions, reverse transcriptase efficiency, polymerase efficiency, etc. We acknowledge that no assay is perfect (ours included), and it is virtually impossible to obtain a “true“ count of an underlying NA concentration. Nevertheless, it is possible to come close using an assay which yields the highest NA concentration, has low background, and produces statistically plausible results. For this, here we have successfully used ultrasensitive digital RT-dPCR. We suggest that RT-dPCR or a similarly improved methodology is implemented for quantification of all RNA stocks used for SARS-CoV-2 assays being submitted for emergency use authorization or equivalent regulatory approval. We also encourage independent evaluations of in vitro diagnostic assay performance using the same quantified standards by unbiased sources.[11]

## Materials and Methods

### Stocks and Dilutions

A DNA plasmid control (2019-nCoV, research-use only) was purchased from Integrated DNA Technologies (IDT; Coralville, Iowa, USA; Cat#148365270, Lot 0000508728; 2×10^5^ copies/µL). Quantitative Synthetic RNA from SARS-Related Coronavirus 2, NR-52358 (Lot 70033953, 5×10^5^ genome equivalents/µL; Lot 70034198, 2.9×10^5^ genome equivalents/µL) and Genomic RNA from SARS-Related Coronavirus 2, Isolate USA-WA1/2020, NR-52285 (Lot 70033700, 5.5×10^4^ genome equivalents/µL; Lot 70033320, 4.8×10^4^ genome equivalents/µL) were obtained from BEI Resources (Manassas, VA, USA). All stocks were aliquoted and stored at -80 C. For all dilution series, an aliquot was thawed and serially diluted in 1x TE buffer with 0.05% Tween-20 in 1.5 mL DNA LoBind Tubes (USA Scientific Incorporated; Ocala, FL, USA).

### RT-qPCR

qPCR was performed using protocol recommendation from the CDC’s Real-Time RT-PCR Panel for Detection of 2019-Novel Coronavirus.[12] Briefly, 5 µL TaqPath 1-Step RT-qPCR Master Mix (ThermoFisher Scientific, Waltham, MA, USA) was added to 1.5 µL combined primer/probe mix (N1; Lot 0000509022 from IDT’s 2019-nCoV CDC qPCR Probe Assay), and 8.5 µL nuclease-free water. Master-mix was added to a 96-well plate (thin-wall clear well, HSP9641, Bio-Rad) and 5 µL of template was mixed by pipette in individual wells. The 96-well plate was sealed (Microseal B, MSB1001, Bio-Rad) and spun briefly in a Mini Plate Spinner Centrifuge (14-100-141, Fisher Scientific) to bring down droplets. Thermocycling and real-time imaging were performed on the Bio-Rad CFX96 Touch Real-Time PCR Detection System (Bio-Rad Laboratories, Hercules, CA, USA) by heating to 25 °C for 2 min, 50 °C for 15 min, 95 °C for 2 min, and cycling 80 times between 95 °C for 3 s and 55 °C for 30 s.

### RT-dPCR

RNA was quantified with reverse transcription droplet digital polymerase chain reaction (RT-dPCR). Reaction mix contained 1X Bio-Rad EvaGreen ddPCR Mix, 200 nM forward and backward COVID primers (N1 primers purchased from IDT, re-suspended in NF-H2O),[13] 1 U/µL Riboguard RNase inhibitor (Lucigen Corp., Madison, WI, USA), and 300 U/mL WarmStart RTx (New England Biolabs, NEB; Ipswich, MA, USA).

Template was added at 10% of the reaction mix and the original concentration calculated from the dilution series. All samples were made to 50 µL and duplicates were run by adding 22 µL to two sample wells in the DG8 Cartridge (1864008, Bio-Rad). Dilutions were quantified using the QX200 droplet digital PCR system (Bio-Rad) and droplet generation, droplet transfer, and foil sealing followed manufacturer’s instructions. Thermocycling took place on a C1000 Touch Thermal Cycler (Bio-Rad) with an RT step at 55 °C for 10 min, pre-melt at 95 °C for 3 min, 40 cycles of 95 °C for 30 s, 60 °C for 30 s, and 68 °C for 30 s, and a stabilization at 4 °C for 5 min, 90 °C for 5 min, and a hold at 12 °C until droplet analysis. A temperature ramp rate of 2 °C/s was used for temperature transitions. Droplets were read according to manufacturer’s instructions. Analysis thresholds were manually set to 10,000. Final concentrations were determined using the merge setting on the QuantaSoft analysis software (Bio-Rad). Measured stock concentrations were calculated by subtracting background (signal average of 16 no-template controls) and multiplying by the dilution factor.

## Data Availability

All data will be made available at CaltechDATA upon publication.

## Conflicts of Interest

R.F.I. receives droplet digital PCR patent royalties from Bio-Rad and has a financial interest in Talis Biomedical Corp.

## Acknowledgements

This work was funded in part by an Innovation in Regulatory Science Award (IRSA) from the Burroughs Wellcome Fund, a grant from the Jacobs Institute for Molecular Engineering for Medicine (Caltech) and NSF Graduate Research Fellowship DGE-144469 (to E.J.). The following reagents were obtained through BEI Resources, NIAID, NIH: Quantitative Synthetic RNA from SARS-Related Coronavirus 2, NR-52358, and Genomic RNA from SARS-Related Coronavirus 2, Isolate USA-WA1/2020, NR-52285. We thank Si Hyung Jin for compiling data from test-kit manufacturer EUAs and Natasha Shelby for contributions to writing and editing this manuscript.

## References

1. CSSE. COVID-19 Dashboard by the Center for Systems Science and Engineering (CSSE) at Johns Hopkins University (JHU) 2020. Available from: https://gisanddata.maps.arcgis.com/apps/opsdashboard/index.html#/bda7594740fd40299423467b48e9ecf6.

2. Johns Hopkins Coronavirus Resource Center. COVID-19 Data Center. Available from: https://coronavirus.jhu.edu/.

3. Federal Register. Determination of Public Health Emergency: A notice by the Health and Human Services Department on 02/07/2020. 2020.

4. FDA. Emergency Use Authorization (EUA) information, and list of all current EUAs 2020. Available from: https://www.fda.gov/emergency-preparedness-and-response/mcm-legal-regulatory-and-policy-framework/emergency-use-authorization#2019-ncov.

5. FDA. Policy for Diagnostic Tests for Coronavirus Disease-2019 during the Public Health Emergency Immediately in Effect Guidance for Clinical Laboratories, Commercial Manufacturers, and Food and Drug Administration Staff. 2020.

6. Sanders R, Huggett JF, Bushell CA, Cowen S, Scott DJ, Foy CA. Evaluation of Digital PCR for Absolute DNA Quantification. Analytical chemistry. 2011;83(17):6474–84. doi: 10.1021/ac103230c.

7. Shen F, Du W, Kreutz JE, Fok A, Ismagilov RF. Digital PCR on a SlipChip. Lab on a chip. 2010;10(20):2666–72. Epub 2010/07/03. doi: 10.1039/c004521g.

8. Majumdar N, Wessel T, Marks J. Digital PCR modeling for maximal sensitivity, dynamic range and measurement precision. PloS one. 2015;10(3):e0118833-e. doi: 10.1371/journal.pone.0118833.

9. Analytical Methods C. Recommendations for the definition, estimation and use of the detection limit. Analyst. 1987;112(2):199–204. doi: 10.1039/AN9871200199.

10. Bio-Rad. Droplet Digital PCR Applications Guide 2014. Available from: http://www.bio-rad.com/webroot/web/pdf/lsr/literature/Bulletin_6407.pdf.

11. FIND. FIND Evaluation update: SARS-CoV-2 Molecular Diagnostics 2020. Available from: https://www.finddx.org/covid-19/sarscov2-eval-molecular/.

12. CDC. Real-Time RT-PCR Panel for Detection 2019-Novel Coronavirus. Centers for Disease Control and Prevention, Respiratory Viruses Branch, Division of Viral Diseases, 2020.

13. CDC. Research Use Only 2019-Novel Coronavirus (2019-nCoV) Real-time RT-PCR Primer and Probe Information 2020. Available from: https://www.cdc.gov/coronavirus/2019-ncov/lab/rt-pcr-panel-primer-probes.html.

